# SARS-CoV-2 CT-scan dataset: A large dataset of real patients CT scans for SARS-CoV-2 identification

**DOI:** 10.1101/2020.04.24.20078584

**Authors:** Eduardo Soares, Plamen Angelov, Sarah Biaso, Michele Higa Froes, Daniel Kanda Abe

**Affiliations:** Lancaster University, School of Computing and Communications, LIRA Research Centre, Lancaster, LA1 4WA, UK; Public Hospital of the Government Employees of Sao Paulo (HSPM), 01532-001, Sao Paulo, BR

## Abstract

The infection by SARS-CoV-2 which causes the COVID-19 disease has widely spread all over the world since the beginning of 2020. On January 30, 2020 the World Health Organization (WHO) declared a global health emergency.Researchers of different disciplines work along with public health officials to understand the SARS-CoV-2 pathogenesis and jointly with the policymakers urgently develop strategies to control the spread of this new disease. Recent findings have observed imaging patterns on computed tomography (CT) for patients infected by SARS-CoV-2. In this paper, we build a public available SARS-CoV-2 CT scan dataset, containing 1252 CT scans that are positive for SARS-CoV-2 infection (COVID-19) and 1230 CT scans for patients non-infected by SARS-CoV-2, 2482 CT scans in total. These data have been collected from real patients in hospitals from Sao Paulo, Brazil. The aim of this dataset is to encourage the research and development of artificial intelligent methods which are able to identify if a person is infected by SARS-CoV-2 through the analysis of his/her CT scans. As baseline result for this dataset we used an eXplainable Deep Learning approach (xDNN) which we could achieve an *F*1 score of 97.31% which is very promising. The proposed dataset is available www.kaggle.com/plameneduardo/sarscov2-ctscan-dataset and xDNN code is available at https://github.com/Plamen-Eduardo/xDNN-SARS-CoV-2-CT-Scan.

## 1 Introduction

In December 2019, an outbreak coronavirus (SARS-CoV-2) infection began in Wuhan, the capital of central China’s Hubei province^1-3^. On January 30, 2020 the World Health Organization (WHO) declared a global health emergency^4^. By 22 April 2020, accumulative 2,564,515 confirmed cases and 177,466 deaths were documented^5^.

Researchers of different disciplines work along with public health officials to understand the COVID-19 pathogenesis and jointly with the policymakers urgently develop strategies to control the spread of this new disease^6^. Recent findings have observed imaging patterns on computed tomography (CT) for patients diagnosed with COVID-19 as prospective analysis revealed bilateral lung opacities on 40 of 41 (98%) chest CTs in infected patients in Wuhan and described lobular and subsegmental areas of consolidation as the most typical findings^6^. Other investigators found high rates of ground-glass opacities and consolidation, sometimes with a rounded morphology and peripheral lung distribution^7,8^. Thoracic radiology evaluation is often key to the evaluation of patients suspected of COVID-19 infection^9^. Prompt detection and diagnosis of the disease is invaluable in the efforts to ensure timely treatment. From a public health perspective, rapid patient isolation is crucial for containment of this communicable disease^4^ and optimal use of available resources which quickly become scarce and overwhelmed by the exponentially growing number of patients and prolonged periods of treatment.

In this paper, we make publicly available a large dataset of CT scans for SARS-CoV-2 identification. The dataset is composed of 1252 CT-scans of patients infected by the SARS-CoV-2 virus and 1230 CT scans of non-infected by SARS-CoV-2 patients, but that have other pulmonary diseases. In order to test the dataset we an eXplainable deep learning method (xDNN)^10^. The eXplainbale approach is non-iterative and is entirely based on recursive calculations and use of prototypes. Therefore, it is computationally very efficient. In this paper we demosntrate that the proposed dataset can be of huge importance for the identification of SARS-CoV-2 via CT scan. Moreover, we demonstrated that the baseline approach, xDNN, can achieve high performance for this challenging task.

## 2 eXplainable Deep Learning (xDNN)

### 2.1 Concept and Basic Algorithm

The prototype-based learning is the core of the xDNN method (Fig. (1)). The prototypes are actual training data samples (in this case, images) which are highly representative (local peaks of the density and empirically derived probability distributions^11^). They are focal points of locally valid generative models described by multi-modal Cauchy distribution^11^.

**Figure 1.**
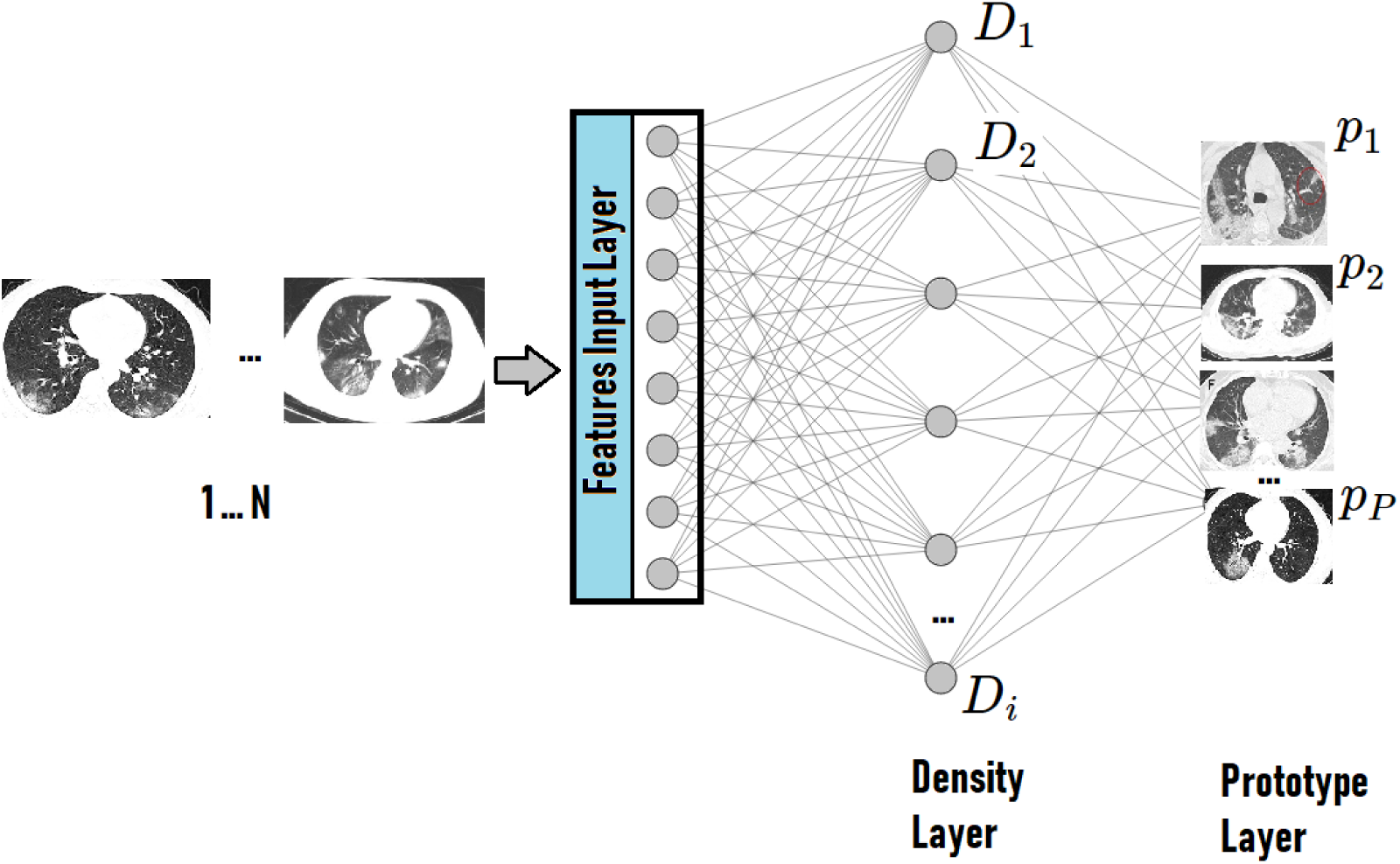
This figure illustrates the layered architecture of the proposed method. It has the form of a deep neural network but is using clear to understand prototypes (actual images). The density layer identifies the local peaks of the density and empirically derived probability distributions. The prototypes are actual training data samples (in this case, images) which are highly representative (local peaks of the density and empirically derived probability distributions.

The algorithm of the proposed approach is described below. With the first observed image (data sample) it is being converted to a vector of features using transfer learning. In this paper, we use a vector with size 4096 formed from the last fully connected layer of the VGG-16^12^.

Let 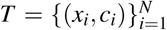 be training data set with *x_i_* ∈ ℝ^n^ denoting the feature vector and *c_i_* ∈ {1,2} denoting the class (SARS-Cov-2 or Non-SARS-Cov-2) for each *i* ∈ {1,…,*N*}. *N* is the number of training data/images used.

The proposed algorithm works per class; therefore, all the calculations are done for each class separately.

The meta-parameters are initialized with the first observed data sample.

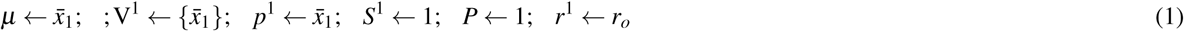

where *μ* denotes the mean; V_1_ denotes the first cluster; *p*_1_ is the first prototype of the first cluster, V_1_; *S*_1_ is the corresponding support (number of members); *P* is the total number of the identified prototypes; *r*_1_ is the corresponding radius of the area of influence of V_1_ (in this paper, we use 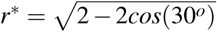 same as^11^; the rationale is that two vectors for which the angle between them is less than π/6 or 30*^o^* are pointing in close/similar directions. That is, we consider that two feature vectors can be considered to be similar if the angle between them is smaller than 30 degrees. Note that *r*^*^ is data derived, not a problem- or user- specific parameter. In fact, it can be defined without *prior* knowledge of the specific problem or data).

The next step is to calculate the data density at the current data point, 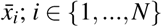.

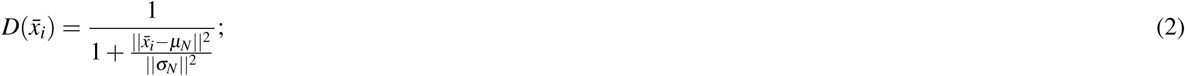

Starting from the mutual distances (Euclidean, Cosine, or Minkowski type) between the data points (samples) in the feature space it can be demonstrated theoretically^11^ that the data density takes the form of a Cauchy type function as in Eq. (2).

Then the algorithm absorbs the new data samples/images, 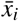 one by one by assigning then to the nearest (in the feature space) prototype, 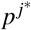:

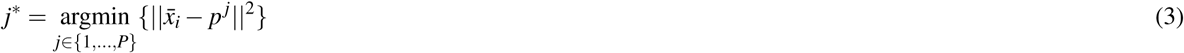

Note that different distance metrics can be used for this type of assignment. Because of this form of assignment, the shape of the data partitioning is of the so-called Voronoi tessellation type^13^. We call all data points associated with a prototype *data clouds*, because their shape is not regular (e.g., hyper-spherical, hyper-ellipsoidal, etc.) and the prototype is not necessarily the statistical and geometric mean^11^.

Then, using the density and the distance to the nearest prototype we check the following conditions^11^ based on which we determine if the current data sample/image is going to be added to the set of prototypes as a new prototype or not:

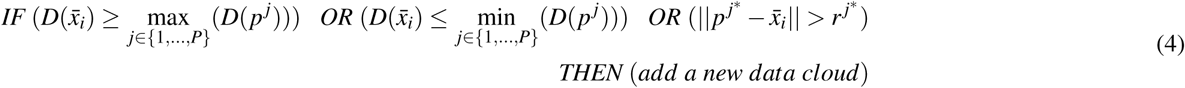

When adding a new data cloud the following updates are being made:

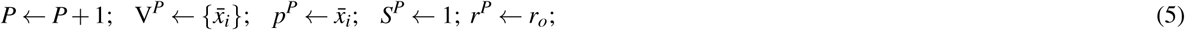

Alternatively, the meta parameters of the nearest data cloud are being updated as follows^11^:

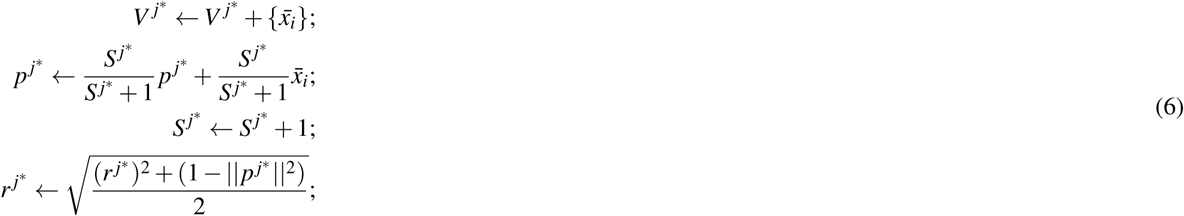

One of the strongest aspects of the proposed approach is its high level of interpretability which comes from its prototype-based nature. Linguistic *IF…THEN* expressions that represent human reasoning can be formed around the local generative models:

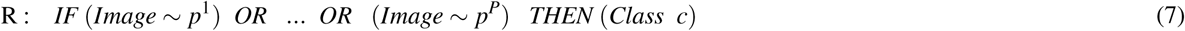

The learning procedure of the proposed approach is summarized by the following algorithm.

#### Learning Procedure

1. Read the first feature vector sample *x_ì_* representing the image *I_i_* of the class *c*;
2. Set 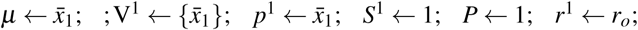
3. **FOR** *i* = 2, …
4. Read 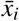;
5. Calculate 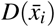 and *D*(*p^j^*) (*j =* 1,*2,..,P)* according to equation (2);
6. **IF** Eq. (4) holds
7. Create rule according to Eq. (5);
8. **ELSE**
9. Search for *p ^j^* according to Eq. (3);
10. Update rule according to Eq. (6);
11. **END**
12. **END**

**Figure 2.**
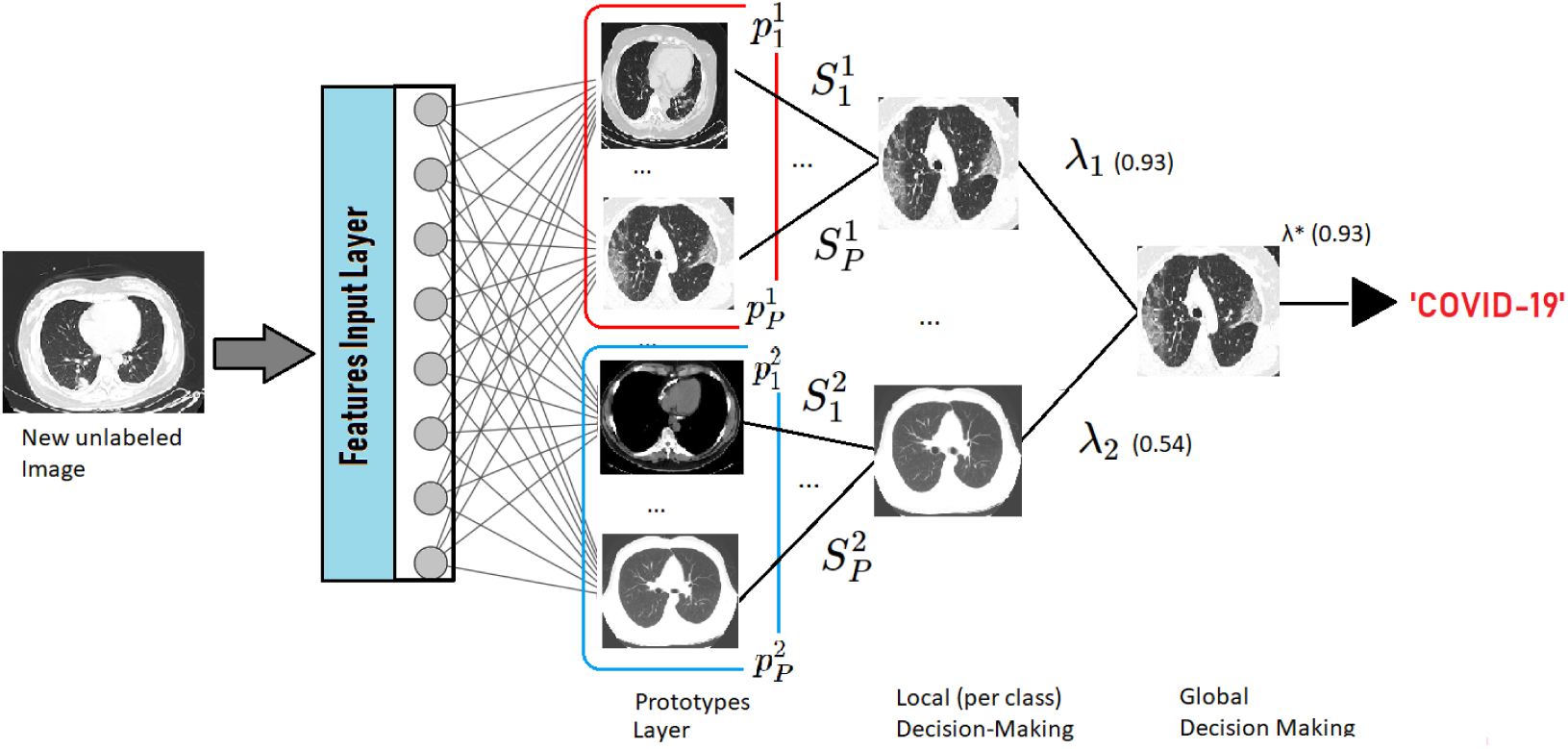
The figure illustrates the decision-making process of the proposed approach. As a new unlabeled data/image arrives it is compared to the identified prototypes. The local (per class) decision-making process is dedicated to calculate the winning prototype per class (the most similar to the new image prototypes of a given class). The global decision-making layer is in charge of forming the overall decision by comparing the degrees of similarity to all classes (in this case, to SARS-CoV-2 or Non-SARS-CoV-2).

## 3 Dataset Description

The proposed dataset is composed of 2482 CT scans images, which is divided between 1252 for patients infected by SARS-CoV-2, and 1230 CT scans for non-infected by SARS-CoV-2 patients, but whom presented other pulmonary diseases. Data was collected from hospitals of Sao Paulo, Brazil. The detailed number of patients is illustrated by Fig (3). The detailed characteristic of each patient has been omitted by the hospitals due to ethical concerns. Fig. (4) illustrates some examples of CT scans for patients infected and non infected by SARS-CoV-2 that composes the dataset.

**Figure 3.**
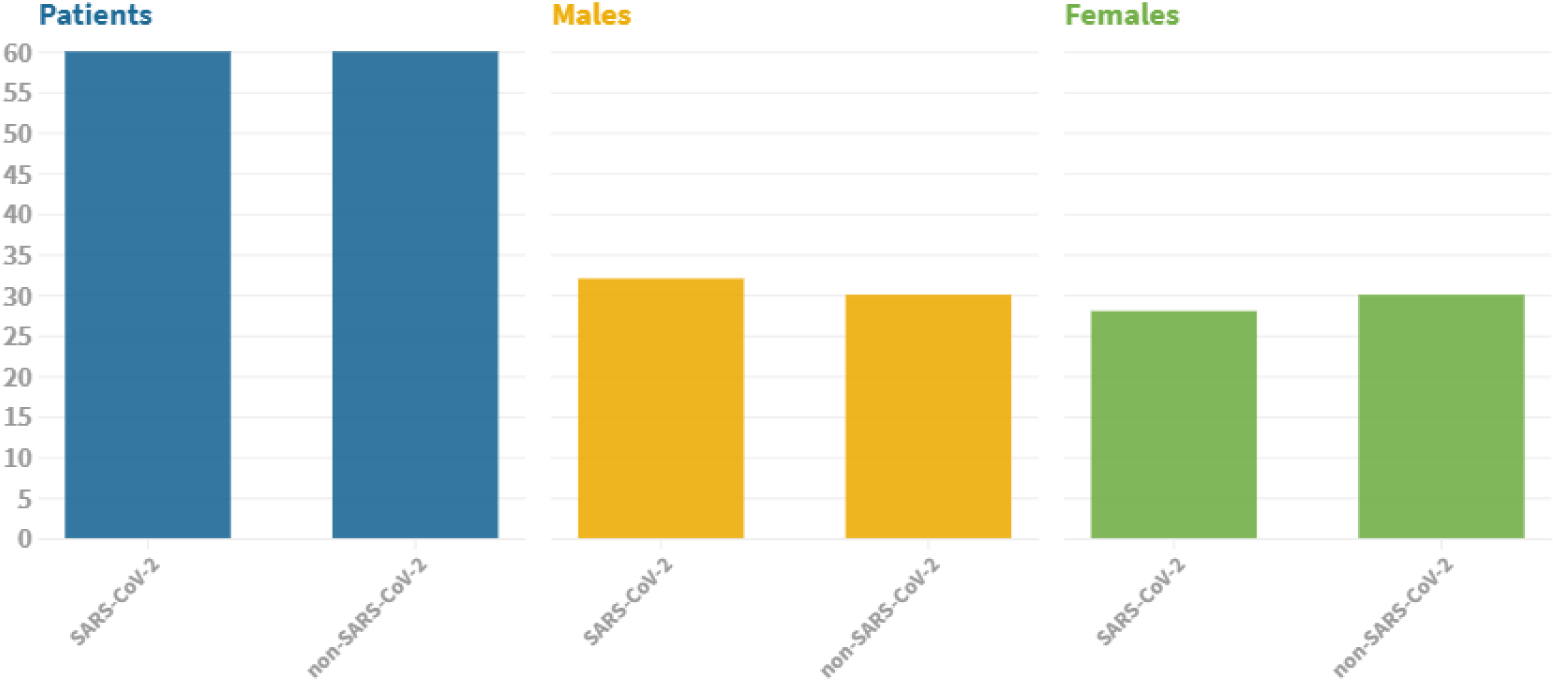
The figure illustrates the number of patients considered to compose the dataset. In this case, we considered data of 60 patients infected by SARS-CoV-2, which 32 are male and 28 are female. We also considered data of 60 patients not infected by SARS-CoV-2, which 30 are male and 30 are female.

**Figure 4.**
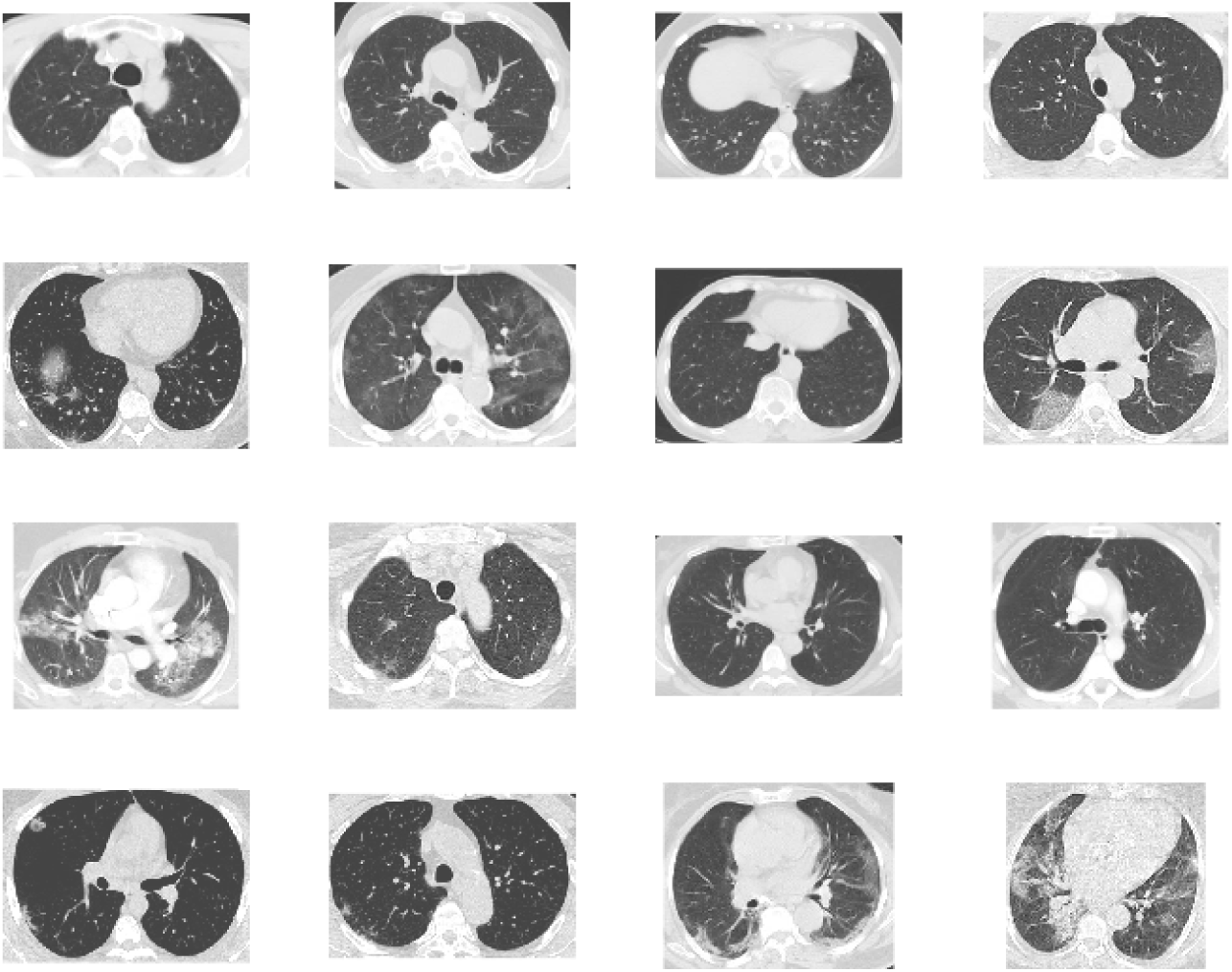
The figure illustrates the example of CT scans for different patients infected and non infected by SARS-CoV-2.

## 4 Results

In this section we report the results obtained by the proposed eXplainable Deep Learning classification approach, xDNN, when applied to the proposed SARS-CoV-2 CT scan dataset. We divided the dataset into 80% for training purposes and 20% for validation purposes. Results presented in Table 1 compare the xDNN algorithm with other state-of-the-art approaches, including traditional (*black-box*) deep neural network, Support vector Machines, etc. In summary, the advantages of the proposed method include:

– high precision as compared with the top state-of-the-art algorithms.
– high level of explainability.
– no user- or problem- specific algorithmic meta parameters
– non-iterative algorithm able to learn continuously.

**Table 1.** The xDNN classifier provided better results in terms of all metrics than the other state-of-the-art approaches, including ResNet, GoogleNet, VGG-16, and Alexnet. Moreover, it also provided highly interpretable results that may be helpful for specialists. Rules generated by the identified prototypes for COVID and Non-COVID patients are illustrated by Figs. (5) and (6) respectively. xDNN spent 11.82 seconds for the training time, an average of 0.005 seconds per image. On the other hand, the traditional deep learninig approach may take hours for the same task.

| Method \ Metric | Accuracy | Precision | Recall | F1 Score | AUC |
| --- | --- | --- | --- | --- | --- |
| xDNN | 97.38% | 99.16% | 95.53% | 97.31% | 97.36% |
| ResNet | 94.96% | 93.00% | 97.15% | 95.03% | 94.98% |
| GoogleNet | 91.73% | 90.20% | 93.50% | 91.82% | 91.79% |
| VGG-16 | 94.96% | 94.02% | 95.43% | 94.97% | 94.96% |
| AlexNet | 93.75% | 94.98% | 92.28% | 93.61% | 93.68% |
| Decision Tree | 79.44% | 76.81% | 83.13% | 79.84% | 79.51% |
| AdaBoost | 95.16% | 93.63% | 96.71% | 95.14% | 95.19% |

Using the proposed method we generated (extracted form the data) linguistic *IF…THEN* rules which involve actual images of both cases (COVID-19 and NO COVID-19) as illustrated in Figs. (5) and (6). Such transparent rules can be used in the decision-making process for early diagnostics for COVID-19 infection. Rapid detection with high sensitivity of viral infection may allow better control of the viral spread. Early diagnosis of COVID-19 is crucial for the disease treatment and control.

**Figure 5.**
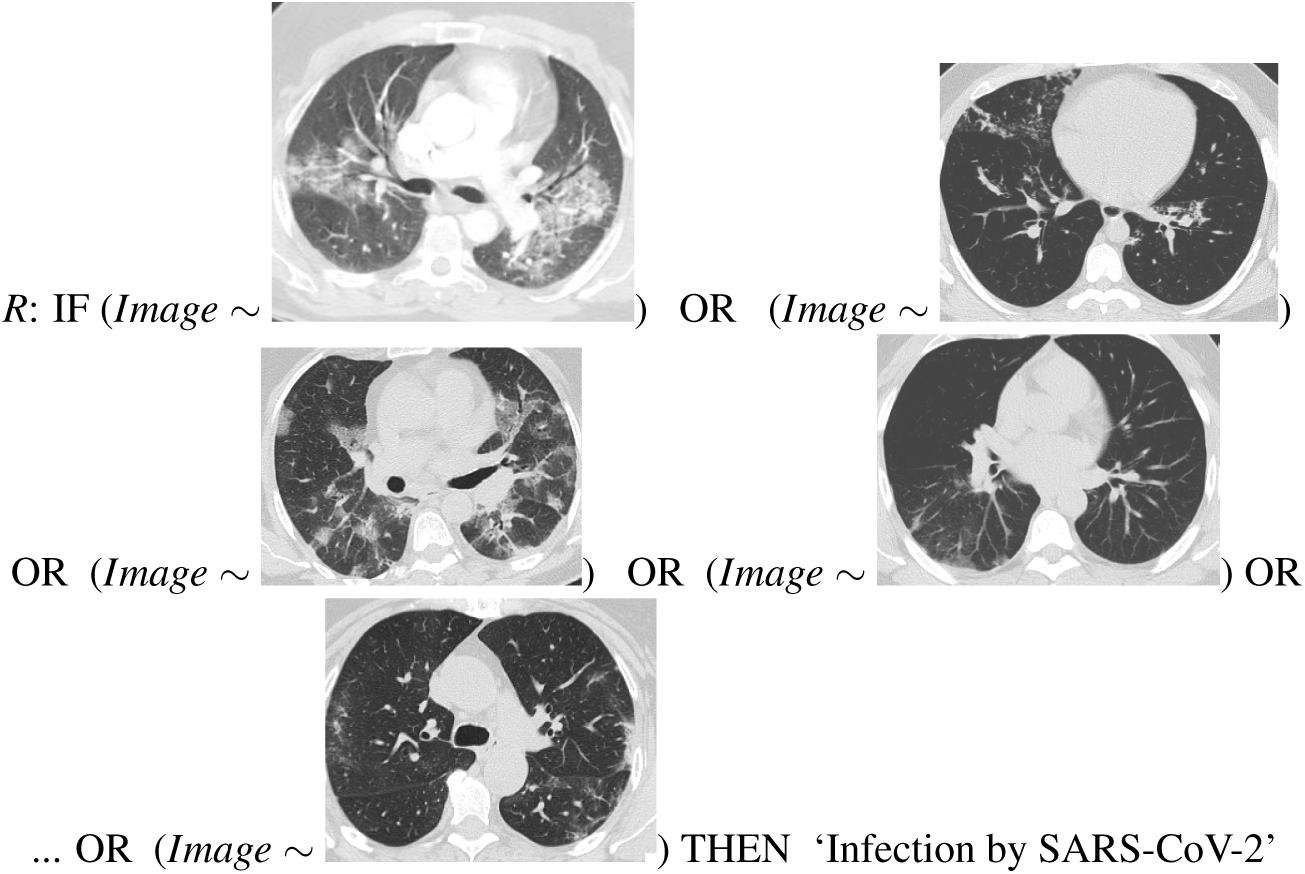
Final rule given by the proposed eXplainable Deep Learning classifier for the COVID-19 identification. Differently from ‘black box’ approaches as deep neural networks, the proposed approach provides highly interpretable rules which can be used by human experts for the early evaluation of patients suspected of COVID-19 infection.

**Figure 6.**
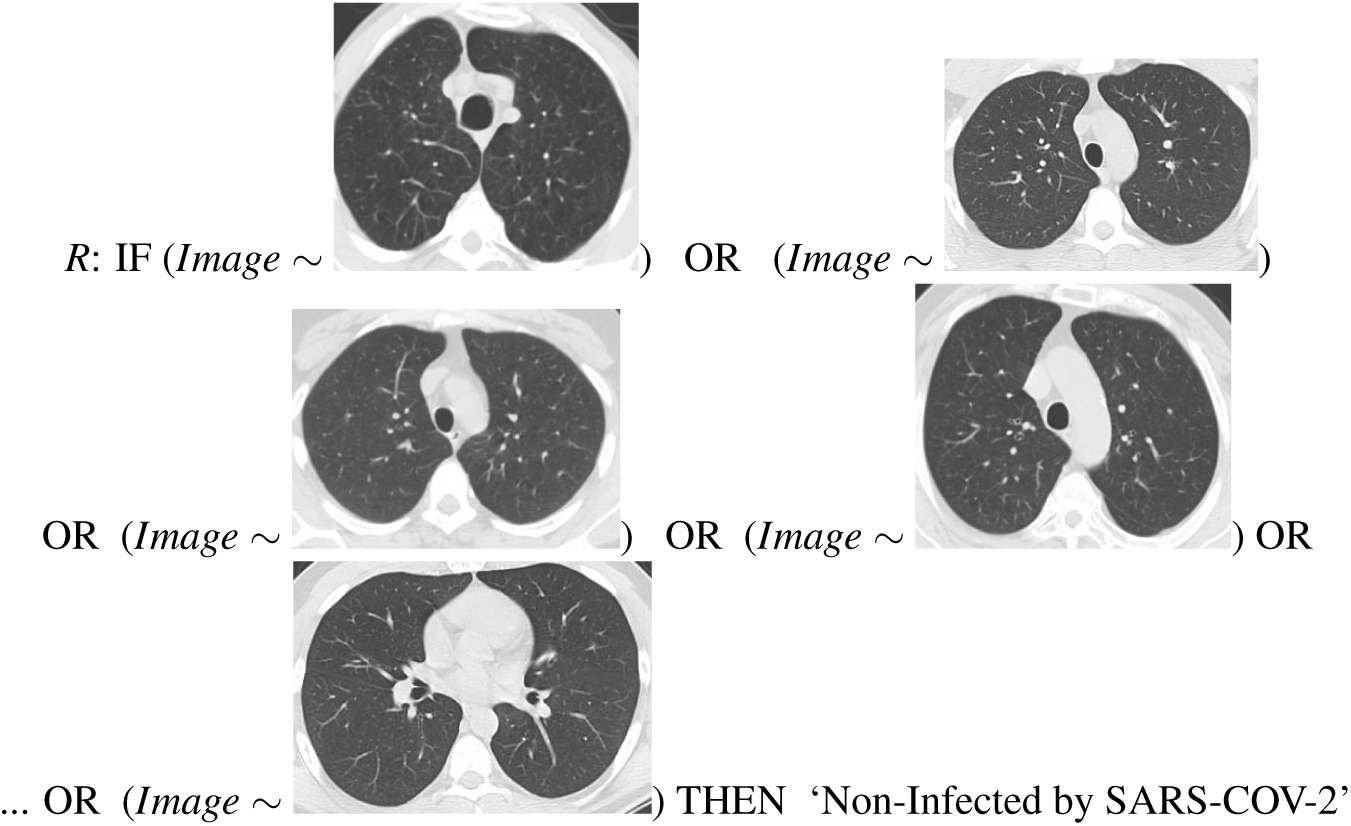
Non-SARS-CoV-2 final rule given by the proposed eXplainable Deep Learning classifier.

Computing tomography is a quick non-invasive imaging modality with high accuracy. According to^14, 15^ almost all patients with COVID-19 had characteristic CT features during the disease, effects such as different degrees of ground-glass opacities with or without crazy-paving sign, multifocal organizing pneumonia, and architectural distortion in a peripheral distribution. The proposed approach has demonstrated high efficiency on the identification and classification of such characteristics, and then provide high accurate and interpretable results.

## 5 Conclusion

In this paper we make public a new large dataset for SARS-CoV-2 identification via CT scans. The presented dataset is composed by 2482 CT scans, which 1252 corresponds to 60 patients identified with SARS-CoV-2 and 1230 CT scans corresponds to 60 patients not identified with SARS-CoV-2. These data has been collected from different hospitals in Sao Paulo, Brazil. Moreover, we tested the proposed dataset has been tested with different methods. The xDNN classifier has presented the best results in terms present a new eXplainable deep learning approach for COVID-19 detection via CT scan. The proposed approach demonstrates better results in terms of performance than other state-of-the-art approaches, presenting an *F*1 score of 97.31% for the best case. Moreover, it also provides epxlanations in the form of *IF…THEN* rules using actual images of CT scans with and without COVID-19. This is of great importance for medical specialists to understand and diagnose COVID-19 at early stages via computed tomography. The proposed dataset is available www.kaggle.com/plameneduardo/sarscov2-ctscan-dataset and xDNN code is available at https://github.com/Plamen-Eduardo/xDNN-SARS-CoV-2-CT-Scan.

## Data Availability

All the data and experiments available in this research paper have been approved by the Ethical Committee of the Public Hospital of the Government Employees of Sao Paulo (HSPM), Sao Paulo/Brazil.

https://www.kaggle.com/plameneduardo/sarscov2-ctscan-datasetand

## Author contributions statement

P. A. conceived and detailed the idea. E. S. designed and implemented the algorithms, designed and performed the experiments.

P. A. and E. S. wrote the manuscript and interpreted the results. E. S., S. B., M. H. F., and D. K. A. collected the data.

## References

1. Huang, C. et al. Clinical features of patients infected with 2019 novel coronavirus in wuhan, china. The Lancet 395, 497–506 (2020).

2. Zhu, N. et al. A novel coronavirus from patients with pneumonia in china, 2019. New Engl. J. Medicine (2020).

3. Zhou, F. et al. Clinical course and risk factors for mortality of adult inpatients with covid-19 in wuhan, china: a retrospective cohort study. The Lancet (2020).

4. Sohrabi, C. et al. World health organization declares global emergency: A review of the 2019 novel coronavirus (covid-19). Int. J. Surg. (2020).

5. Dong, E., Du, H. & Gardner, L. An interactive web-based dashboard to track covid-19 in real time. The Lancet infectious diseases (2020).

6. Ai, T. et al. Correlation of chest ct and rt-pcr testing in coronavirus disease 2019 (covid-19) in china: a report of 1014 cases. Radiology 200642 (2020).

7. Ng, M.-Y. et al. Imaging profile of the covid-19 infection: radiologic findings and literature review. Radiol. Cardiothorac. Imaging 2, e200034 (2020).

8. Kong, W. & Agarwal, P. P. Chest imaging appearance of covid-19 infection. Radiol. Cardiothorac. Imaging 2, e200028 (2020).

9. Shi, H. et al. Radiological findings from 81 patients with covid-19 pneumonia in wuhan, china: a descriptive study. The Lancet Infect. Dis. (2020).

10. Angelov, P. & Soares, E. Towards eXplainable deep neural networks (xdnn). *arXivpreprint arXiv:1912.02523* (2019).

11. Angelov, P. P., & Gu, X. Empirical approach to machine learning (Springer, 2019).

12. Simonyan, K. & Zisserman, A. Very deep convolutional networks for large-scale image recognition. *arXiv preprint arXiv:1409.1556* (2014).

13. Du, Q., Faber, V. & Gunzburger, M. Centroidal voronoi tessellations: Applications and algorithms. SIAM review 41, 637–676 (1999).

14. Bernheim, A. et al. Chest ct findings in coronavirus disease-19 (covid-19): relationship to duration of infection. Radiology 200463 (2020).

15. Zhao, W., Zhong, Z., Xie, X., Yu, Q. & Liu, J. Relation between chest ct findings and clinical conditions of coronavirus disease (covid-19) pneumonia: a multicenter study. Am. J. Roentgenol. 1–6 (2020).

